# Rapid adaptation of established high-throughput molecular testing infrastructure for detection of monkeypoxvirus

**DOI:** 10.1101/2022.06.05.22276011

**Authors:** Dominik Nörz, Hui Ting Tang, Petra Emmerich, Katja Giersch, Nicole Fischer, Marylyn M. Addo, Martin Aepfelbacher, Susanne Pfefferle, Marc Lütgehetmann

**Affiliations:** University Medical Center Hamburg-Eppendorf (UKE), Institute of Medical Microbiology, Virology and Hygiene, Hamburg, Germany; Bernhard-Nocht institute for tropical medicine (BNITM), Molecular Biology and Immunology, Virology Department, Hamburg, Germany; University Medical Center Hamburg-Eppendorf (UKE), Institute for Infection research and Vaccine development (IIRVD), Hamburg, Germany; Bernhard-Nocht Institute for Tropical Medicine (BNI), Department of Clinical Immunology of Infectious Diseases, Hamburg, Germany; German Center for Infection Research, partner site Hamburg-Lübeck-Borstel-Riems

## Abstract

**Background:** Since May 2022, a rising number of monkeypox-cases has been reported in non-endemic countries of the northern hemisphere. In contrast to previous clusters, infections seem predominantly driven by human-to-human transmission, rather than animal sources. In this study, we adapted two published qPCR assays (non-variola orthopoxvirus and monkeypoxvirus specific) for use as a lab-developed dual-target monkeypoxvirus-test on widely used automated high-throughput PCR-systems (cobas5800/6800/8800).

**Methods:** Selected assays were checked for *in-silico* inclusivity and exclusivity in current orthopoxvirus sequences, as well as for multiplex compatibility. Analytic performance was determined by serial dilution of monkeypoxvirus reference material, quantified by digital PCR. Cross reactivity was ruled out through a clinical exclusivity set containing various bloodborne and respiratory pathogens. Clinical performance was compared to a commercial manual RUO-kit using clinical remnant samples.

**Results:** Analytic lower limit of detection (LoD) was determined as 4.795 dcp/ml (CI95%: 3.598 - 8.633 dcp/ml) for both assays combined, with a dynamic range of at least 5 log-steps. The assay showed 100% positive and negative agreement with the manual RUO orthopoxvirus PCR test kit in clinical swab samples.

**Discussion:** While the full extend of the ongoing monkeypox outbreak remains to be established, the WHO and local health authorities are calling for increased awareness and efforts to limit further spread. For this, timely and scalable PCR tests are an important prerequisite. The assay presented here allows streamlined high-throughput molecular testing for monkeypoxvirus on existing hardware, broadly established previously for SARS-CoV-2 diagnostics.

## 1. Introduction

In May 2022, an unusually high number of monkeypox cases were reported in Western European and North American countries (257 lab confirmed infections as of 29/05/22), including Spain, Portugal, the United Kingdom, Canada and the United States, sparking the fear of another global outbreak (1). Endemic transmission of the monkeypox virus (a species of the orthopox genus) is thought to be limited to Central and Western Africa, with both zoonotic (est. 22% - 72% of cases) and person-to-person transmission playing significant roles for the burden of disease in these areas (2). Clusters outside Africa have usually been traceable to animal sources in the past, rather than human-to-human transmission (3). In contrast, the current cases seem to have occurred without any links to animal sources and are concentrated in, but not exclusive to, the MSM (male having sex with male) community (4). The sudden appearance of infections in several non-endemic countries suggests that undetected transmission may have taken place for some time, but certain recent events may have served as a catalyst for spread (1).

The ongoing SARS-CoV-2 pandemic has demonstrated the potential and importance of highly automated high-throughput molecular testing in outbreak scenarios. The aim of this study was to rapidly adapt the existing automated molecular testing infrastructure for SARS-CoV-2 in a large tertiary care hospital (i.e. cobas5800/6800/8800 platforms) for detection of monkeypoxvirus from clinical samples, thereby creating the capacity for high-throughput testing and quick turn-around times if needed.

## 2. Material and Methods

### 2.1 Multiplex assay setup (NVAR/MPOX/IC)

Based on recent experiences with diagnostics during the SARS-CoV-2 outbreak (5, 6), a dual-target approach was chosen, with one assay targeting a conserved sequence of the orthopoxvirus genus, not including variola major/minor (Li et al, viral gene: E9L; Target-1: “NVAR” (7)) and the other a sequence specific for monkeypoxvirus (Shchelkunov et al. viral gene: B7R, Target-2: “MPOX” (8)). The cobas 5800/6800/8800 systems use a spike-in RNA full-process control, added automatically during extraction. The corresponding internal-control assay is preloaded in the open channel reagent (MMX-R2, cobas omni Utility Channel).

All assays were modified and optimized for use on the cobas5800/6800/8800 systems, including 2’O-methyl-RNA-modified primers and internal quenchers for taqman-probes, as previously described (9).

**Table 1:**
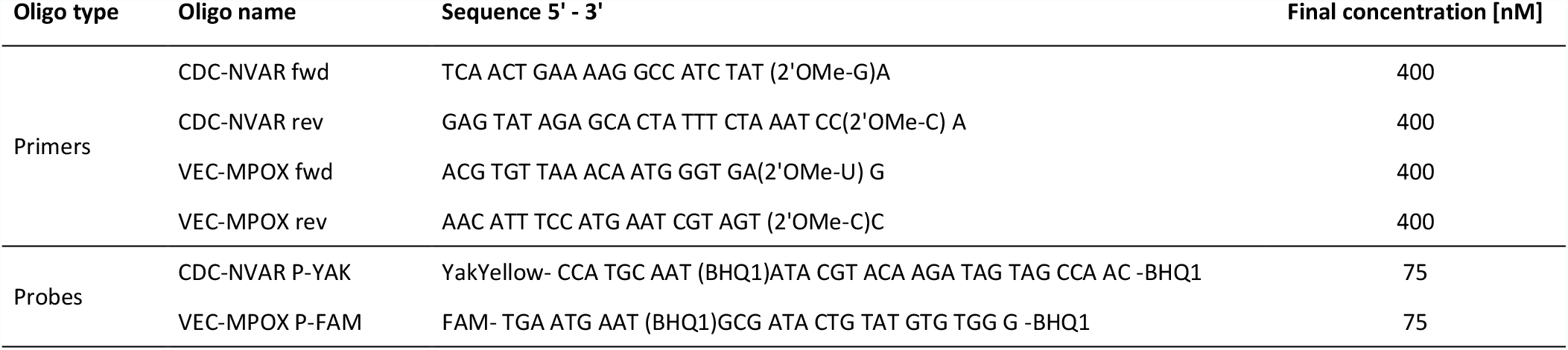
Primer-, probe- sequences of the duplex assay are listed. Oligos were custom made by Ella Biotech GmbH (Fürstenfeldbruck, Germany). Indicated final concentration refer to the final oligo concentrations within the reaction mix. 2’O-methyl-RNA bases are indicated as “OMe-X”. Internal abasic quenchers are indicated as (BHQ1)”).

**Table 2:** Cobas omni Utility Channel run protocol for the NVAR_MPOX-UCT. RFI (relative fluorescence increase) thresholds are used for automated result calls.

| Software settings |  |  |  |  |  |
| --- | --- | --- | --- | --- | --- |
| Sample type | Swab (400 µL) |  |  |  |  |
| Channels | 1: Not used | 2: MPOX | 3: NVAR | 4: Not used | 5: IC |
| RFI |  | 2 | 2 |  | 2 |
| PCR cycling conditions |  |  |  |  |  |
|  | UNG incubation | Pre-PCR step | 1<sup>st</sup> measurement | 2<sup>nd</sup> measurement | Cooling |
| No. of cycles | Predefined | 1 | 5 | 45 | Predefined |
| No. of steps |  | 3 | 2 | 2 |  |
| Temperature |  | 55°C; 60°C; 65°C | 95°C; 55°C | 91°C; 58°C |  |
| Hold time |  | 120 s; 360 s; 240 s | 5 s; 30 s | 5 s; 25 s |  |
| Data acquisition |  | None | End of each cycle | End of each cycle |  |

### 2.2 In-silico evaluation

As part of a support request for Utility channel applications, all sequences of the multiplex-assay were submitted to Roche Diagnostics (Pleasanton, CA) for evaluation of inclusivity and potential primer/probe interactions. Assay sequences were aligned to currently available Monkeypoxvirus and Orthopoxvirus sequences available in public databases.

### 2.3 Evaluation of analytical performance

Technical performance evaluation for the assay was performed according to new EU regulations (2017/746 EU IVDR). Reference material used for this study was inactivated cell culture supernatant containing monkeypoxvirus, recovered from a clinical case in central Africa in 1987 (10), and inactivated MVA SARS-CoV-2 vaccine (Vacciniavirus, Ankara strain derived). To obtain a quantitative monkeypoxvirus standard, nucleic acids were purified using a MagNA-pure96 extractor (Roche diagnostics, Rotkreuz, Switzerland) and analysed on a QIAcuity digital PCR instrument (Qiagen, Hilden, Germany) in conjunction with three different qPCR assays (NVAR by Li. Et al, MPXV by Shchelkunov et al. and LightMix Modular Orthopoxvirus by Tibmol-biol (RUO), Berlin, Germany).

Lower limit of detection (LoD) was determined by serial two-fold dilution of monkeypoxvirus standard in universal transport medium (UTM) from 100 digital copies (dcp)/ml to 0.78 dcp/ml, 21 repeats per dilution step (prepared using a Hamilton IVD STARlet liquid handler, Hamilton, Bonaduz Switzerland). Linearity was assessed by 10-fold serial dilution of Monkeypoxvirus standard (5 repeats per dilution step) between concentrations of approximately 10^1^ dcp/ml and 10^7^ dcp/ml, analyzed. Linearity and Intra-assay variability were determined using Validation Manager software (Finbiosoft, Espoo, Finland). For empirical inclusivity/exclusivity testing, a set of clinical samples, reference material and external quality controls of a range of bloodborne and respiratory pathogens was tested with the assay (53 samples in total). An experimental MVA vector-based SARS-CoV-2 vaccine was used as reference material for a non-monkeypox orthopoxvirus.

### 2.4 Clinical evaluation

For clinical validation, the LightMix Modular Orthopoxvirus assay (RUO; TibMol-biol, Berlin, Germany) was used as reference test, which was performed according to manufacturer’s recommendation using the MagNA-pure96 system with 200µl extraction volume. In total, 72 clinical samples were tested with both assays, consisting of respiratory, skin and genital swabs. Of these, six samples were positive for monkeypoxvirus-DNA, obtained from two current clinical cases in Hamburg, Germany.

## 3. Results

### 3.1 in-silico analysis

There were no concerning oligo interactions (*see supplementary figure 1*). Target-1: NVAR is still a 100% match for all but a single monkeypoxvirus sequence (with one low-risk mismatch). NVAR also has high sequence similarity with many other orthopoxviruses, but may not be optimal for reliable detection of e.g. camelpox or cowpox (*see supplementary figure 2*). Target-2: MPOX is a perfect match for almost all congo-basin-strain monkeypoxvirus sequences, but has a known mismatch for west-Africa-strain sequences in the probe region. This is expected to slightly reduce relative fluorescence increase (RFI)-signals, as demonstrated in the clinical sample set (see below). Other orthopoxviruses have extensive sequence mismatches with this assay and are not expected to produce detectable signals (*see supplementary figure 3*).

### 3.2 Analytical performance

LoD was determined as 9.697 dcp/ml (CI95%: 7.424 – 15.327 dcp/ml) for the NVAR-assay and 6.359 dcp/ml (CI95%: 4.908 – 10.110 dcp/ml) for the MPOX-assay by probit analysis using medcalc software (i.e. 95% probability of detection). Overall LoD (both targets combined) was 4.795 dcp/ml (CI95%: 3.598 - 8.633 dcp/ml). Probit plots are available in *supplementary figure 4*.

**Table 3:** LoDs were determined by serial dilution of a quantified monkeypoxvirus standard (quantified by digital PCR) as reference. Dilution series were generated automatically using a Hamilton STARlet IVD liquid handler. 95% probability of detection was calculated using medcalc software.

| NVAR_MPOX-UCT |  |  |  |
| --- | --- | --- | --- |
| Conc. [dcp/ml] | NVAR | MPOX | Overall |
| 100 | 21/21 | 21/21 | 21/21 |
| 50 | 21/21 | 21/21 | 21/21 |
| 25 | 21/21 | 21/21 | 21/21 |
| 12.5 | 20/21 | 21/21 | 21/21 |
| 6.25 | 19/21 | 20/21 | 21/21 |
| 3.125 | 11/21 | 13/21 | 15/21 |
| 1.56 | 11/21 | 11/21 | 17/21 |
| 0.78 | 3/21 | 6/21 | 7/21 |

The assay showed excellent linearity between ct37 and ct18, approximately 10^1^ dcp/ml and 10^7^ dcp/ml, with pooled SD and CI within linear range: 0.194 Ct and 0.0662% and 0.175 Ct and 0,618% for (NVAR and MPOX respectively). (See also *figure 1*).

**Figure 1:**
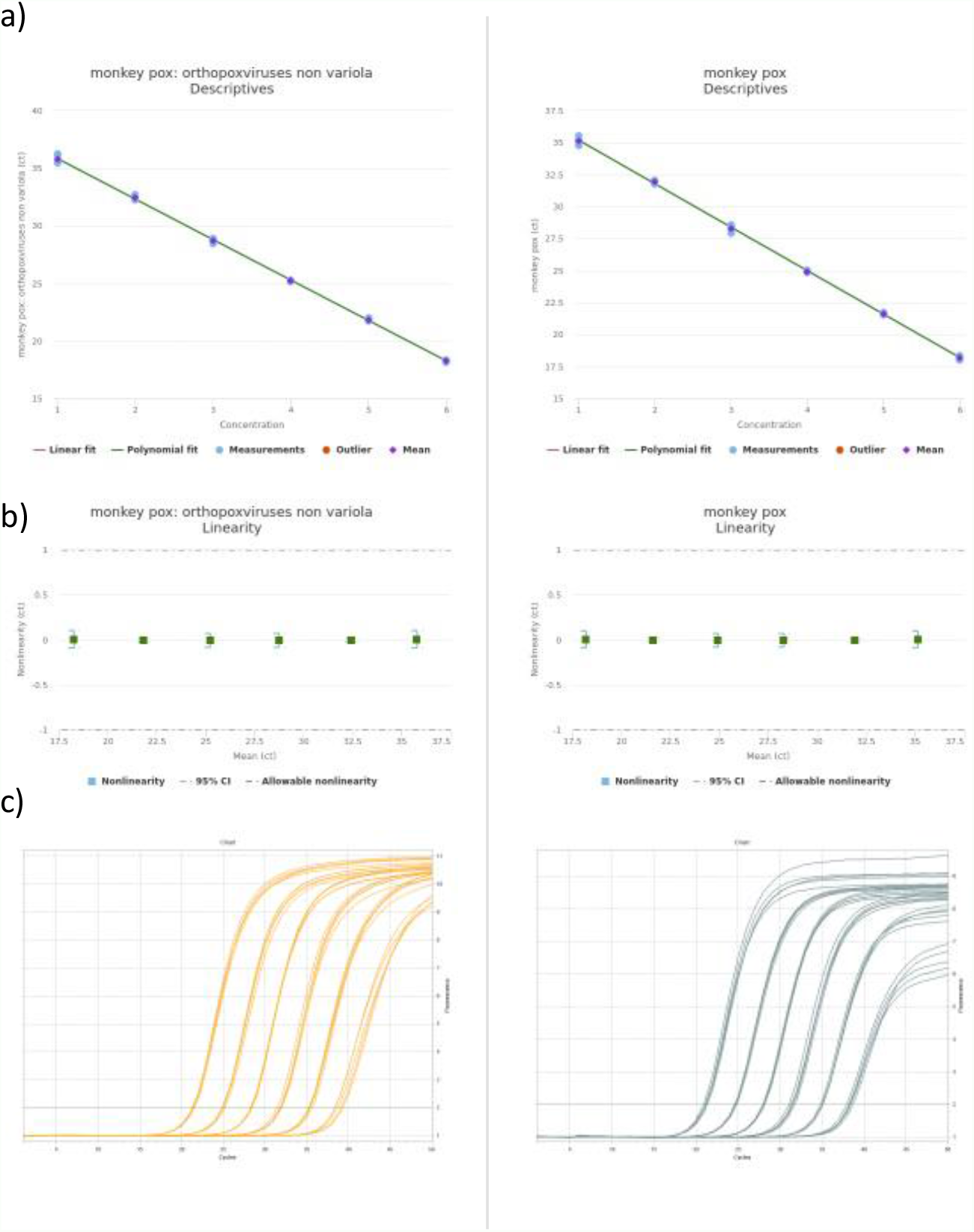
Linearity data for Target-1: NVAR (left) and Target-2: MPOX (right). Linearity was determined by serial dilution of monkeypoxvirus reference material (cell culture supernatant, 1987). Analysis was carried out using Validationmanager software (Finbiosoft, Espoo, Finland). Target-1: NVAR: slope -3.52, r^2^0.999; Target-2: MPOX: slope -3.40, r^2^: 0.999)

No false positives occurred within the inclusivity/exclusivity set. The MVA vector vaccine was correctly detected by the NVAR assay, and not by the MPOX assay (see *supplementary table 1*).

### 3.3 Clinical evaluation

In total, 72 clinical samples were tested with both assays, consisting of respiratory, skin and genital swabs. Of these, six samples were positive for monkeypoxvirus-DNA, obtained from two current clinical cases in Hamburg, Germany. There was a 100% positive (6/6) and 100% negative agreement (56/56). (See figure 2 for amplification curves of positive patient samples)

**Figure 2:**
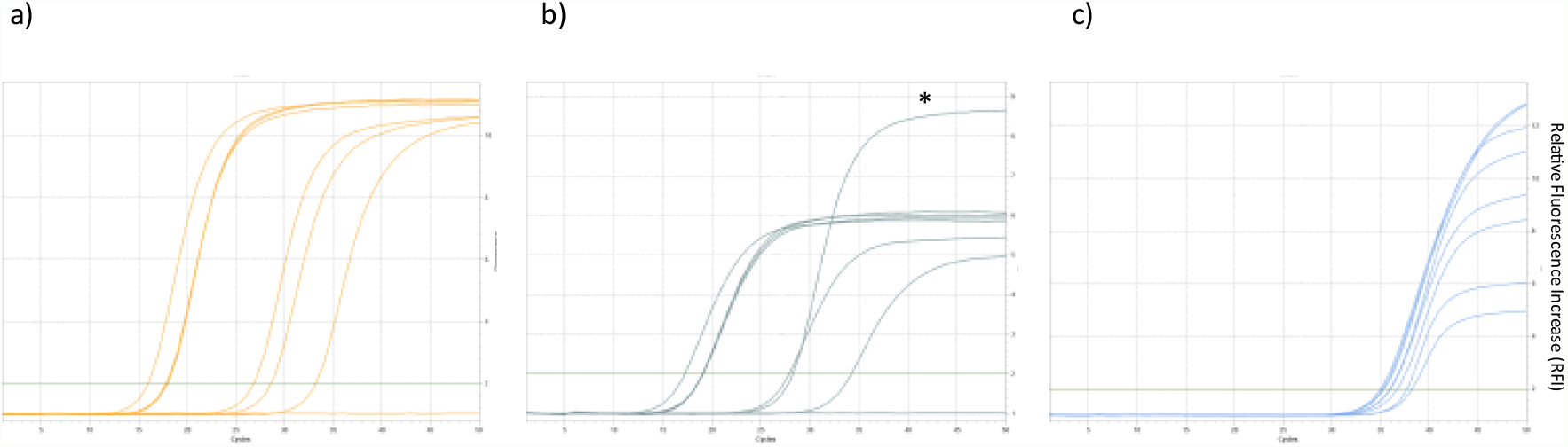
Amplification curves of the clinical sample set for a) Target-1: NVAR, b) Target-2: MPOX, c) internal control. Samples were swabs of lesions, oropharyngeal swabs and EDTA plasma. * indicates the positive control curve in Channel 2 (i.e. cell culture supernatant of Congo basin strain, 1987). West Africa strain samples exhibit an approx. 1/3 reduction in RFI for Target-2: MPOX, due to a known mismatch in the probe region. (See supplementary figure 1).

## 4. Discussion and Conclusion

Significant uncertainties remain around the trajectory of the ongoing monkeypoxvirus outbreak in Europe and North America. However, the WHO acknowledges that these current clusters represent a change in pattern and emphasizes the need to limit further spread (1), for which broad availability of molecular testing with short turn-around times is a crucial prerequisite.

In this study we adapted two established qPCR assays as a duplex test for the cobas5800/6800/8800 fully automated sample-to-result platforms, which are widely used for high-throughput SARS-CoV-2 diagnostics (11). Both assays have been validated extensively against other Orthopoxvirus species by the respective authors (7, 8) and remain inclusive and highly specific in *in-silico* analysis with currently available monkeypox sequences. We demonstrated excellent analytical performance of the duplex-assay, with single digit detection limits and near perfect PCR efficiency. A spike-in full-process control assay, similar to commercial CE-IVD assays, is already included in the open channel reagents.

During the preparation of this manuscript, two clinical cases of monkeypox were confirmed by our institution, the clinical samples of which constitute the entirety of our clinical positive-set. The low number of individual patients and clinical samples represents a limitation of this study. Though the assay was only validated on swab samples, monkeypoxvirus-DNA was also detectable in EDTA plasma in relevant concentrations without any adaptations of the method. Generally, the quantitative ratios of viral DNA in different sample types were very well in line with recently published data (swab from lesions: ct15, oropharyngeal swab: ct24, blood: ct30) (12). Further studies may be needed to evaluate the practical usefulness of plasma samples for diagnostic purposes or longitudinal viral-load monitoring.

In conclusion, we provided technical performance validation for a lab-developed duplex qPCR-assay for monkeypoxvirus detection on the cobas5800/6800/8800 high-throughput systems. The assay allows adaptation of existing automated molecular testing infrastructure from the SARS-CoV-2 pandemic for a potential large-scale monkeypox outbreak.

## Data Availability

All data produced in the present study are available upon reasonable request to the authors

## 6. Abbreviations

Dcp: digital copies
LoD: Limit of Detection
IC: internal control
IVD: in-vitro diagnostic
RFI: relative fluorescence increase
CI: confidence interval

## 7. Author contribution

ML, SP and DN conceptualized and supervised the study. HTT, DN, PE, performed the experiments. KG performed data analysis. DN, ML, SP, NF, MAd and MAe wrote and edited the manuscript. All authors agreed to the publication of the final manuscript.

## References

1. WHO. 2022. Disease Outbreak News; Multi-country monkeypox outbreak in non-endemic countries (29 May 2022).

2. Bunge EM, Hoet B, Chen L, Lienert F, Weidenthaler H, Baer LR, Steffen R. 2022. The changing epidemiology of human monkeypox—A potential threat? A systematic review. PLOS Neglected Tropical Diseases 16:e0010141.

3. Reed KD, Melski JW, Graham MB, Regnery RL, Sotir MJ, Wegner MV, Kazmierczak JJ, Stratman EJ, Li Y, Fairley JA, Swain GR, Olson VA, Sargent EK, Kehl SC, Frace MA, Kline R, Foldy SL, Davis JP, Damon IK. 2004. The Detection of Monkeypox in Humans in the Western Hemisphere. New England Journal of Medicine 350:342–350.

4. Adalja A, Inglesby T. 2022. A Novel International Monkeypox Outbreak. Annals of Internal Medicine doi:10.7326/M22-1581.

5. Corman VM, Landt O, Kaiser M, Molenkamp R, Meijer A, Chu DK, Bleicker T, Brunink S, Schneider J, Schmidt ML, Mulders DG, Haagmans BL, van der Veer B, van den Brink S, Wijsman L, Goderski G, Romette JL, Ellis J, Zambon M, Peiris M, Goossens H, Reusken C, Koopmans MP, Drosten C. 2020. Detection of 2019 novel coronavirus (2019-nCoV) by real-time RT-PCR. Euro Surveill 25.

6. Manohar C, Sun J, Schlag P, Santini C, Fontecha M, Lötscher P, Bier C, Goepfert K, Duncan D, Spier G, Jarem D, Kosarikov D. 2021. Agile design and development of a high throughput cobas^®^ SARS-CoV-2 RT-PCR diagnostic test. medRxiv doi:10.1101/2021.10.13.21264919:2021.10.13.21264919.

7. Li Y, Olson VA, Laue T, Laker MT, Damon IK. 2006. Detection of monkeypox virus with real-time PCR assays. Journal of Clinical Virology 36:194–203.

8. Shchelkunov SN, Shcherbakov DN, Maksyutov RA, Gavrilova EV. 2011. Species-specific identification of variola, monkeypox, cowpox, and vaccinia viruses by multiplex real-time PCR assay. Journal of Virological Methods 175:163–169.

9. Pfefferle S, Reucher S, Nörz D, Lütgehetmann M. 2020. Evaluation of a quantitative RT-PCR assay for the detection of the emerging coronavirus SARS-CoV-2 using a high throughput system. Eurosurveillance 25:2000152.

10. Müller G, Meyer A, Gras F, Emmerich P, Kolakowski T, Esposito JJ. 1988. MONKEYPOX VIRUS IN LIVER AND SPLEEN OF CHILD IN GABON. The Lancet 331:769–770.

11. Poljak M, Kor a M, Knap Gašper N, ujs Komloš K, Sagadin M, Uršič T A, šič Županc T, Petroec M. 2020. Clinical evaluation of the cobas SARS-CoV-2 test and a diagnostic platform switch during 48 hours in the midst of the COVID-19 pandemic. Journal of Clinical Microbiology doi:10.1128/jcm.00599-20:JCM.00599-20.

12. Adler H, Gould S, Hine P, Snell LB, Wong W, Houlihan CF, Osborne JC, Rampling T, Beadsworth MBJ, Duncan CJA, Dunning J, Fletcher TE, Hunter ER, Jacobs M, Khoo SH, Newsholme W, Porter D, Porter RJ, Ratcliffe L, Schmid ML, Semple MG, Tunbridge AJ, Wingfield T, Price NM, Abouyannis M, Al-Balushi A, Aston S, Ball R, Beeching NJ, Blanchard TJ, Carlin F, Davies G, Gillespie A, Hicks SR, Hoyle M-C, Ilozue C, Mair L, Marshall S, Neary A, Nsutebu E, Parker S, Ryan H, Turtle L, Smith C, van Aartsen J, Walker NF, Woolley S, Chawla A, Hart I, Smielewska A, et al. Clinical features and management of human monkeypox: a retrospective observational study in the UK. The Lancet Infectious Diseases doi:10.1016/S1473-3099(22)00228-6.

