## Supplemental material for "Rapid adaptation of established high-throughput molecular testing infrastructure for detection of monkeypoxvirus"

Supplementary figure 1

#### Potential oligo-oligo interaction

1:

```
                GAGTATAGAGCACTATTTCTAAATCCCACACATACAGTATCGCATTCATTCA
CDC-NVAR rev    ............................
                                        ............................    VEC-MPOX P-FAM
```

*The CDC-NVAR reverse primer and VEC-MPOX probe show a 3′ overlap of 4 bases. While the interaction cannot create dimers on its own, similar interactions can in rare cases create a viable dimer in a secondary unspecific amplification event, thereby also leading to unspecific signals. The risk for such an event is further mitigated by 2′O-methyl-RNA modified primers and internal quenchers. No unspecific signals were observed in wetlab experiments. Unspecific signals of the MPOX assay would yield non-sensical results (i.e. NVAR negativ, MPOX positive), prompting further investigation. Based on these factors, the interaction was deemed not significant.*

Supplementary figure 2

#### Target-1: NVAR

| group | n (Sequences) | hits | forward-1(f)<br>TCAACTGAAAAGGCCATCTATGA | probe-1(p)<br>CCATGCAATATACGTACAAGATAGTAGCCAAC | reverse-1(r)<br>GAGTATAGAGCACTATTTCTAAATCCCA |
|---|---|---|---|---|---|
| Monkeypox | **n = 105** | 1 | ....................... | ................................ | ............................ |
| Monkeypox | n = 1 | 1 | .................T..... | ................................ | ............................ |
| Abatino_macacapox | n = 2 | 1 | ....................... | ................................ | ..............G............. |
| Akhmeta | n = 5 | 1 | ..C.................... | ................................ | ..............G............. |
| Akhmeta | n = 2 | 1 | ..C........A........... | .............A.....A..C......... | ............................ |
| Camelpox | n = 10 | 1 | ....................... | .......G..............C......... | ............................ |
| Cowpox | n = 29 | 1 | ....................... | ................................ | ............................ |
| Cowpox | n = 2 | 1 | ....................... | ......................C......... | ........C..G................ |
| Cowpox | n = 5 | 1 | ....................... | ................................ | ........C..G................ |
| Cowpox | n = 1 | 1 | ....................... | .......G..............C......... | ...........G................ |
| Cowpox | n = 1 | 1 | ....................... | .......G..............C..G...... | ............................ |
| Cowpox | n = 5 | 1 | ....................... | ................................ | ..............G............. |
| Cowpox | n = 1 | 1 | ....................... | ......................C......... | ..............G............. |
| Cowpox | n = 2 | 1 | ..............T........ | .......G..............C......... | ............................ |
| Cowpox | n = 5 | 1 | ....................... | ................................ | ........C..G................ |
| Ectromelia | n = 13 | 1 | ....................... | ................................ | ..............G............. |
| Raccoonpox | n = 5 | 1 | ..T.................... | .............A.....A.....T..G..T | .....G...................... |
| Skunkpox | n = 4 | 1 | ..T........A........... | .............T..T..A........G..T | .....G...................... |
| Taterapox | n = 3 | 1 | ....................... | .......G..............C......... | ............................ |
| Vaccinia | n = 31 | 1 | .................T..... | ................................ | ............................ |
| Vaccinia | n = 1 | 1 | ......A................ | ..........G................A.... | A........................... |
| Vaccinia | n = 72 | 1 | ....................... | ................................ | ............................ |
| Vaccinia | n = 11 | 1 | .................T..... | ...................A............ | ............................ |
| Vaccinia | n = 11 | 1 | ....................... | ...................A............ | ............................ |
| Variola | n = 57 | 1 | ....................... | .......G..............CA........ | ............................ |
| Volepox | n = 4 | 1 | ..T........A.....A..... | .A...........T..T..A........G..T | .....G...................... |

*Alignments were provided by Roche Diagnostics (Pleasanton, USA) as part of a support request. Currently available Orthopoxvirus sequences were checked for missmatches with the NVAR assay. The number of sequences represened by each line is indicated on the left. There exists a single monkeypovirus sequence with 1 primer missmatch, with a low risk of relevant impact.*

#### Supplementary figure 3

##### Target-2: MPOX

| group | id/number | hits | forward-1(f) ACGTGTTAAACAATGGGTGATG | probe-1(p) TGAATGAATGCGA---T---A---CTGTATGTGTGGG | reverse-1(r) AACATTTCCATGAATCGTAGTCC |
|---|---|---|---|---|---|
| Monkeypox | **n = 1** | 1 | ............T......... | .............---.---.---............. | ....................... |
| Monkeypox | **n = 22** | 1 | ...................... | .........T...---.---.---............. | ....................... |
| Monkeypox | **n = 77** | 1 | ...................... | .............---.---.---............. | ....................... |
| Abatino_macacapox | n = 2 | 1 | .....................A | ........CA...---.---GTTT.......C..... | ...............A...T... |
| Camelpox | n = 10 | 1 | .T.....C.............A | ........CA...---.---GTTG..........A.. | ..............CA....... |
| Cowpox | n = 8 | 1 | .......C.............A | ........C..A.---.---GTTG..........A.. | ..............C........ |
| Cowpox | n = 24 | 1 | .......C.............A | ........CA...---.---GTTG..........A.. | ..............C........ |
| Cowpox | n = 1 | 1 | .....................A | ........CA...---.---GTTG............. | ....................... |
| Cowpox | n = 4 | 1 | .......C.............A | ........C..A.---.---GTTG..........T.. | ..............C........ |
| Cowpox | n = 2 | 1 | G....................A | ........CA...---.---GTTG............. | ....................... |
| Cowpox | n = 1 | 1 | T......C.............A | ........CA...---.---GTTG..........A.. | ..............C........ |
| Cowpox | n = 2 | 1 | ...................... | ........C....---.---.TTA............. | ....................... |
| Raccoonpox | n = 3 | 1 | .........G........A..T | .T..C....A...CGT.---.---.C.....C..A.. | .................C.A... |
| Skunkpox | n = 2 | 1 | ...AA....G........A..C | .A.......A...---.GTCG---.....G....A.. | ...................A... |
| Taterapox | n = 2 | 1 | .......C.............A | ........CA...---.---GTTG..........A.. | ..............C........ |
| Vaccinia | n = 1 | 1 | ...................... | ........C..A.---.---GTTT.....A.C..T.. | ...............A...T... |
| Vaccinia | n = 1 | 1 | ...................... | ........C..A.---.---.TTA............. | ....................... |
| Vaccinia | n = 8 | 1 | ...................... | ........C....---.---.TTA............. | ....................... |
| Vaccinia | n = 10 | 1 | ...................... | ........C....---.---GTTT.....A.C..T.. | ....................... |
| Vaccinia | n = 73 | 1 | ...................... | ........C....---.---GTTT.....A.C..T.. | ...............A...T... |
| Vaccinia | n = 28 | 1 | ...................... | ........C....---.---.TTA............. | ...............A...T... |
| Vaccinia | n = 1 | 1 | ..........T....A...... | ..T.....C..T.---.---.TTA............. | ....................... |
| Vaccinia | n = 1 | 1 | .............A........ | ........C....---.---GTTT.....A.C..T.. | .....A.........A...T... |
| Vaccinia | n = 1 | 1 | ...................... | ........C....---.---GTTT.....A.C..T.. | ....................... |
| Variola | n = 76 | 1 | .......T.............A | CATCATT....C.---.TA-.---G.....C.A.CAC | ..............CA....... |
| Variola | n = 2 | 1 | .......C.............A | ........CA...---.---GTTG..........A.. | ..............C........ |
| Variola | n = 2 | 1 | .......C.............A | ........CA...---.GTTG---.-.........A. | ..............C........ |
| Variola | n = 2 | 1 | .......C.............A | ........CA...---.---G---T..-C.....A.. | ..............CA....... |
| Volepox | n = 2 | 1 | ...AA....G........C..C | .T..C....A...---.GTT.---..........A.. | .................C.A.A. |

*Alignments were provided by Roche Diagnostics (Pleasanton, USA) as part of a support request. Currently available Orthopoxvirus sequences were checked for missmatches with the MPOX assay. The number of sequences represened by each line is indicated on the left. Monkeypovirus strains of the West Africa strain have a known missmatch in the Probe region (G to T), leading to a reduction in RFI of about 1/3. (See figure 2)*

### Supplementary figure 4

#### Target-1: NVAR

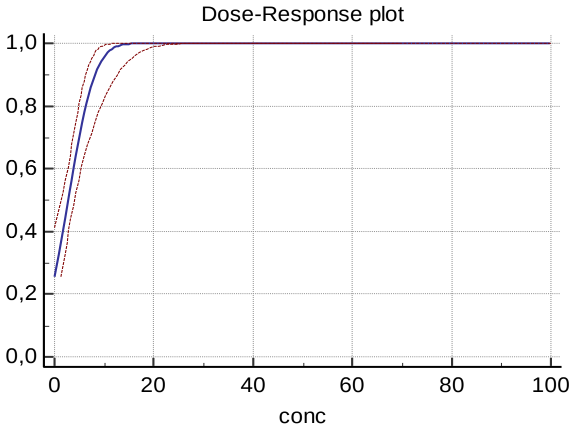


#### Target-2: MPOX

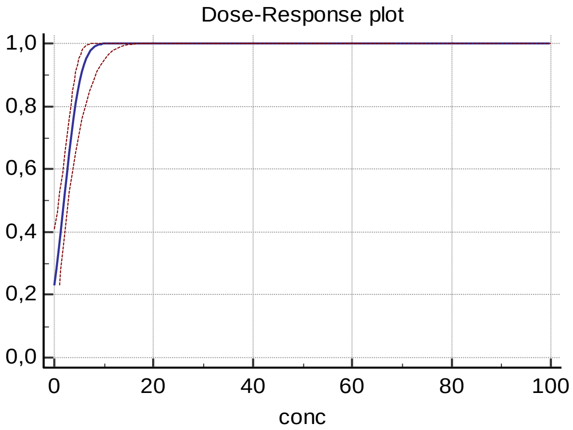


#### Target-1/2 combined

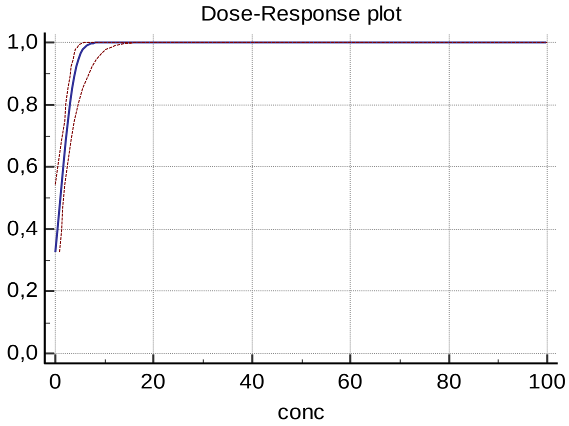


*Probit curves of the LoD (lower Limit of Detection) experiment. Briefly, a 2-fold dilution series of quantified monkeypoxirus standard (quantified by digital PCR) was used to determine the 95% probability of detection (21 repeats per dilution step). Confidence intervals are indicated in red. Hit-rates are displayed in table 3.*

### Supplementary table 1

| External Ref. Material, Quality Control Panel | | | |
|---|---|---|---|
| Species | Number tested | Target: NVAR | Target: MPOX |
| Monkeypoxvirus (Congo basin, cell culture, 1987) | 3 | **Positive** | **Positive** |
| MVA-SARS-2-ST_HD (Vacciniavirus, Ankara strain) | 1 | **Positive** | Negative |
| Influenza-B (INSTAND e.V.) | 1 | Negative | Negative |
| Parainfluenzavirus-1 (Zeptometrix) | 1 | Negative | Negative |
| Parainfluenzavirus-4 (Zeptometrix) | 1 | Negative | Negative |
| Human Coronavirus OC43 (Zeptometrix) | 1 | Negative | Negative |
| Clinical samples | | | |
| Species | | Target: NVAR | Target: MPOX |
| HSV-1 | 5 | Negative | Negative |
| VZV | 4 | Negative | Negative |
| CMV | 5 | Negative | Negative |
| EBV | 5 | Negative | Negative |
| HHV6 | 5 | Negative | Negative |
| BK-Virus | 5 | Negative | Negative |
| JC-Virus | 4 | Negative | Negative |
| SARS-CoV-2 | 1 | Negative | Negative |
| Influenza-A | 1 | Negative | Negative |
| Respiratory syncytial virus | 1 | Negative | Negative |
| Parainfluenzavirus-3 | 1 | Negative | Negative |
| Rhino-/Enterovirus | 3 | Negative | Negative |
| Adenovirus | 2 | Negative | Negative |
| Bocavirus | 2 | Negative | Negative |
| Human Coronavirus HKU1 | 1 | Negative | Negative |

*Clinial samples, reference material and external control panels used for the inclusivity/exclusivity set (53 in total). No false positives occured. The MVA-SARS-2 Vacciniavirus based vaccine (Ankara strain) was correctly detected by the NVAR assay and not by the MPOX assay.*
